# Evaluation of performance of two SARS-CoV-2 Rapid whole-blood finger-stick IgM-IgG Combined Antibody Tests

**DOI:** 10.1101/2020.05.27.20112888

**Authors:** Thierry Prazuck, Mathilda Colin, Susanna Giachè, Camélia Gubavu, Aymeric Seve, Vincent Rzepecki, Marie Chevereau-Choquet, Catherine Kiani, Victor Rodot, Elsa Lionnet, Laura Courtellemont, Jérôme Guinard, Gilles Pialoux, Laurent Hocqueloux

**Author notes:** **Corresponding Author** Thierry Prazuck, Service des maladies infectieuses, CHR Orléans, 14 avenue de l’hôpital, 45 067 Orléans, France.

## Abstract

**Background:** The SARS-CoV-2 virus is responsible for the infectious respiratory disease called COVID-19 (COronaVIrus Disease). In response to the growing COVID-19 pandemic, Rapid Diagnostic Tests (RDTs) have been developed to detect specific antibodies, IgG and IgM, to SARS-CoV-2 virus in human whole blood. We conducted a real-life study to evaluate the performance of two RDTs, COVID-PRESTO^®^ and COVID-DUO^®^, compared to the gold standard, RT-PCR.

**Methods:** RT-PCR testing of SARS-Cov-2 was performed from nasopharyngeal swab specimens collected in adult patients visiting the infectious disease department at the hospital (Orléans, France). Fingertip whole blood samples taken at different time points after onset of the disease were tested with RDTs. The specificity and sensitivity of the rapid test kits compared to test of reference (RT-PCR) were calculated.

**Results:** Among 381 patients with symptoms of COVID-19 who went to the hospital for a diagnostic, 143 patients were RT-PCR negative. Results of test with RDTs were all negative for these patients, indicating a specificity of 100% for both RDTs.

In the RT-PCR positive subgroup (n=238), 133 patients were tested with COVID-PRESTO^®^ and 129 patients were tested with COVID-DUO^®^ (24 patients tested with both). The further the onset of symptoms was from the date of collection, the greater the sensitivity. The sensitivity of COVID-PRESTO^®^ test ranged from 10.00% for patients having experienced their 1^st^ symptoms from 0 to 5 days ago to 100% in patients where symptoms had occurred more than 15 days before the date of tests. For COVID-DUO^®^ test, the sensitivity ranged from 35.71% [0-5 days] to 100% (> 15 days).

**Conclusion:** COVID-PRESTO^®^ and DUO^®^ RDTs turned out to be very specific (none false positive) and to be sensitive enough after 15 days from onset of symptom. These easy to use IgG/IgM combined test kits are the first ones allowing a screening with capillary blood sample, by typing from a finger prick. These rapid tests are particularly interesting for screening in low resource settings.

## Introduction

At the end of 2019, a pneumonia of unknown cause detected in Wuhan, China was first reported to the WHO Country Office in China. On January 9^th^, 2020, the Chinese health authorities and the World Health Organization (WHO) officially announced the discovery of a novel coronavirus, first named 2019-nCoV, then officially termed SARS-CoV-2. This virus, belonging to the coronavirus family, differs from the viruses SARS-CoV, responsible for the SARS outbreak in 2003, and MERS-CoV, responsible for an ongoing outbreak that began in 2012 in the Middle East.

The SARS-CoV-2 virus is responsible for the infectious respiratory disease called COVID-19 (COronaVIrus Disease). This infection mainly results in pneumonia and upper/lower respiratory tract infection. The symptoms of COVID-19 infection appear after an incubation period of approximately 5.2 days [1]. The most common symptoms at onset of COVID-19 illness are fever, cough, and fatigue, but others include headache, sore throat, and even acute respiratory distress syndrome, leading to respiratory failure.

Since the emergence of COVID-19 in China at the end of last year, the SARS-CoV-2 virus has caused a large global outbreak and has become a major worldwide public health issue. The WHO has declared this outbreak a global health emergency at the end of January 2020. On April 12^th^, 2020, the World Health Organization (WHO) announced that the total global deaths from COVID-19 has surpassed 100 000. Globally, by April 28^th^, 2020, 2,892,688 cases of COVID-19 have been confirmed and 210,193 patients have died. An estimated 1.7 billion people have been ordered to remain at home as governments take extreme measures to protect their populations.

Due to the rapid spread and increasing number of COVID-19 cases caused by this new coronavirus SARS-CoV-2, rapid and accurate detection of virus and/or disease is increasingly vital to control the sources of infection and prevent the progression of the disease.

Besides the main priority, which is finding an efficient treatment, one of the most important research questions targets the diagnosis of COVID-19. Currently, the real-time RT-PCR assay is the gold-standard method to detect SARS-CoV-2 [2]. This diagnostic test aims at detecting nucleic acid (RNA) from SARS-CoV-2 in upper and lower respiratory specimens such as nasopharyngeal or oropharyngeal swabs or broncho-alveolar lavage.

In response to the growing COVID-19 pandemic, serology tests have been developed to detect specific antibodies, IgG and IgM, to SARS-CoV-2 virus in human whole blood, serum or plasma. Two kinds of serologic tests are currently available [3]: quantitative ones with antibodies titration by enzyme-linked immunosorbent assay (ELISA) and qualitative ones with Rapid Diagnostic Tests (RDTs), easy-to-use devices mainly based on lateral flow chromatographic immunoassays.

COVID-PRESTO^®^ and COVID-DUO^®^ are two RDTs products with CE marking which are marketed by AAZ-LMB. In line with the recommendations of the health authorities, we conducted a real-life study to evaluate the performance of both AAZ COVID 19 IgM/IgG RDTs compared to the gold standard, RT-PCR.

## Methods and Materials

### Ethics Approval

The study was approved by the local Ethics Committee on March 17^th^ 2020, and informed consent was obtained from each participant.

### Study population

The study population consisted of adult patients visiting the infectious disease department (Centre Hospitalier Regional Orléans, France) from March, 18^th^, 2020 to April 10^th^, 2020. This department receives patients whose symptoms, such as headache, fatigue, fever or respiratory signs suggest a COVID infection, and for whom a diagnosis is requested. Date of onset of symptoms as declared by the patient and age were collected at inclusion. According to severity of disease, patients RT-PCR positive were either hospitalized in the infectious diseases ward, only devoted to treat COVID-19 infected patients, or invited to have regular medical visits in the outpatient consultation. Fingertip blood samples were performed at various stages of the follow-up, even after clinical cure, in order to collect samples from convalescent patients.

### Specimen collection

Nasopharyngeal (NP) swab specimens were collected from patients by trained surveillance officers. A polyester-tipped flexible aluminum-shafted applicator (Microtest M4RT, Remel) was inserted into one of the nostrils until resistance was felt at the nasopharynx, then rotated 180 degrees and withdrawn. After swabbing, the swab applicator was cut off, and each absorbent swab was placed into a vial containing 3 mL of viral transport media. Vials were immediately shipped via a triple packaging system to the virology unit located in the same building of the hospital, then stored if necessary at 4°C for up to 24 hours until testing.

For whole blood samples taken at the fingertip, a lancet was used to prick the side of the fingertip to let a large drop of suspended blood form. This blood sample was collected with a 10 μl capillary micropipette that filled automatically. The sample was then expelled by squeezing the micropipette bulb to deposit the blood on the appropriate well of the test cassette. Retesting was performed in a same patient only if the previous test was negative.

### Real-time RT-PCR assays for the detection of SARS-CoV-2

RT-PCR testing of SARS-CoV-2 was performed in Unit of Virology, CHR Orléans. Nucleic acid extraction was performed with automated EZ1 (Qiagen). Specific real-time RT-PCR assays targeting two RNA-dependent RNA polymerases (IP2 and IP4) and E genes were used to detect the presence of SARS-CoV-2 following the instructions in the protocols of the Institut Pasteur and Corman et al., respectively [4] [5]. Amplification was performed on an ABI 7900 Sequence Detection System (Applied Biosystem).

### Rapid diagnostic tests to be assessed

The SARS-CoV-2 IgG/IgM antibody test kits, COVID-PRESTO^®^ and COVID-DUO^®^, are targeting on the antibodies specific to N-protein of SARS-CoV-2. They are manufactured and marketed by AAZ-LMB.

Tests were conducted at the site by clinical staff, physicians or nurses, according to manufacturers’ instructions. Health workers involved in the study received a two-hours training session for each type of test prior to the beginning of the study.

Both COVID-PRESTO^®^ and COVID-DUO^®^ are lateral flow immune-chromatographic assays (Figs 1 and 2). These tests use anti-human IgM antibody (test line IgM), anti-human IgG antibody (test line IgG) and rabbit IgG (control line C) immobilized on a nitrocellulose strip. The Conjugate (recombinant COVID-19 antigens labeled with colloidal gold) is also integrated into the strip. When a specimen is added to the sample well, followed by assay buffer, IgM and IgG antibodies, if present, will bind to COVID-19 conjugates forming an antigen-antibodies complex.

**Fig 1.**
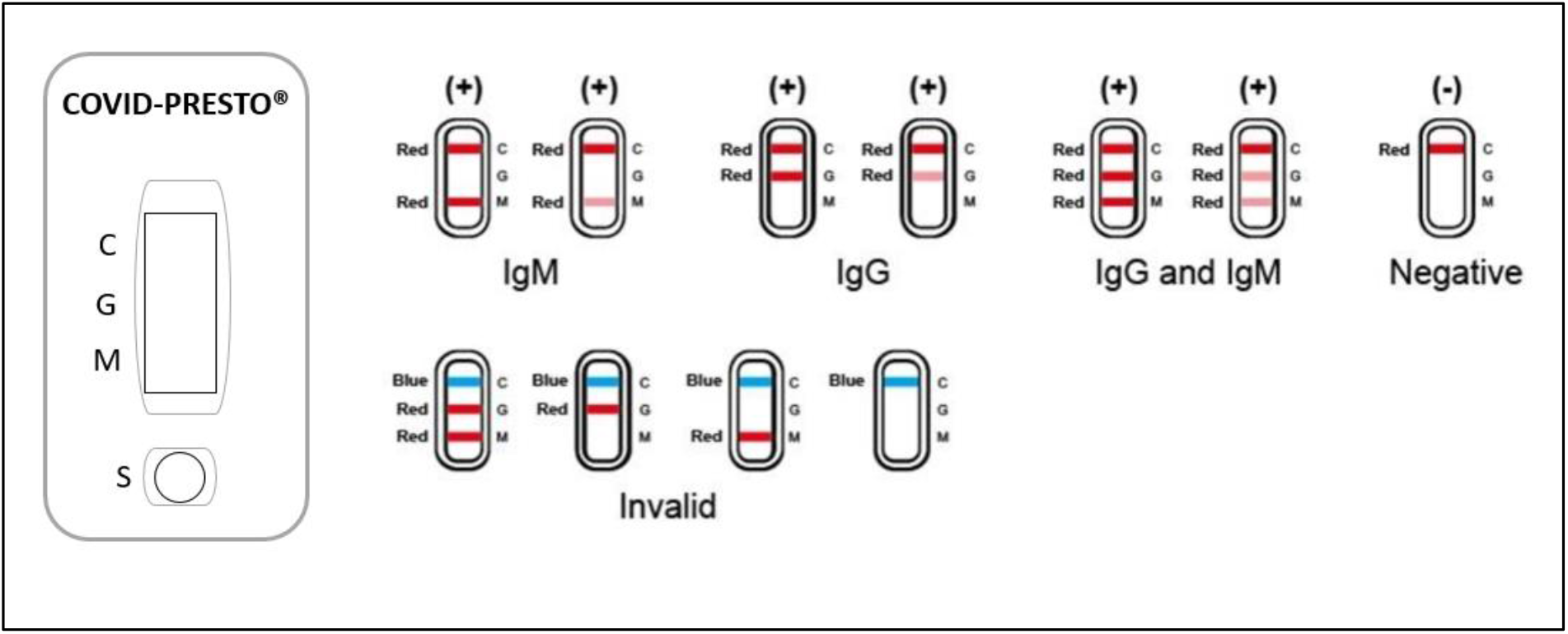
Interpretation of results for COVID-PRESTO^®^.

**Fig 2.**
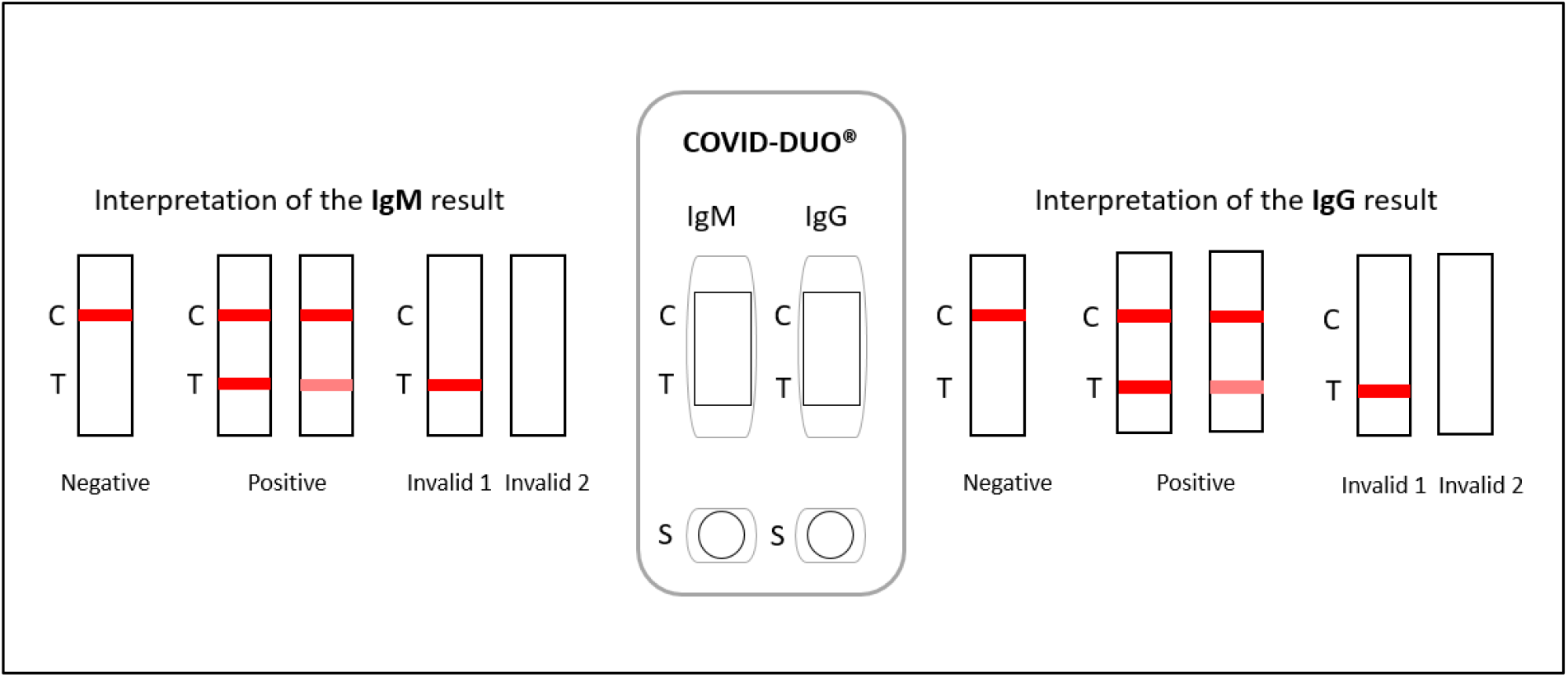
Interpretation of results for COVID-DUO^®^.

This complex migrates through nitrocellulose membrane by capillary action. When the complex meets the line of the corresponding immobilized antibody (anti-human IgM and/or anti-human IgG), the complex is trapped, forming a burgundy colored band which confirms a reactive test result. The result has to be read within 10 minutes by two independent operators. When the control line is the only to be burgundy, the sample is negative. If the control line does not appear, the test is invalid and should be repeated with a new cassette.

### Data Analysis

Population were described in terms of %, mean, standard deviation, range and median values.

The test data was analyzed in the Department of Infectiology. The specificity and sensitivity of the rapid test kits compared to test of reference (RT-PCR) were calculated according to the following formulas:

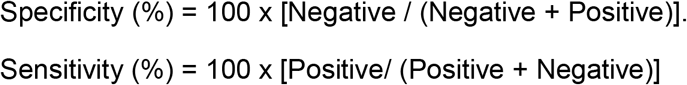

Confidence intervals for sensitivity were produced with the Wilson score method [6].

## RESULTS

Overall, 381 patients with symptoms of COVID-19 who went to the hospital for a diagnostic, were included in the study.

RT-PCR was performed in all patients: 62.47% were positive (n=238). Based on these results, two sub-groups were defined: 143 patients with negative and 238 patients with positive RT-PCR results (Fig 3).

**Fig 3.**
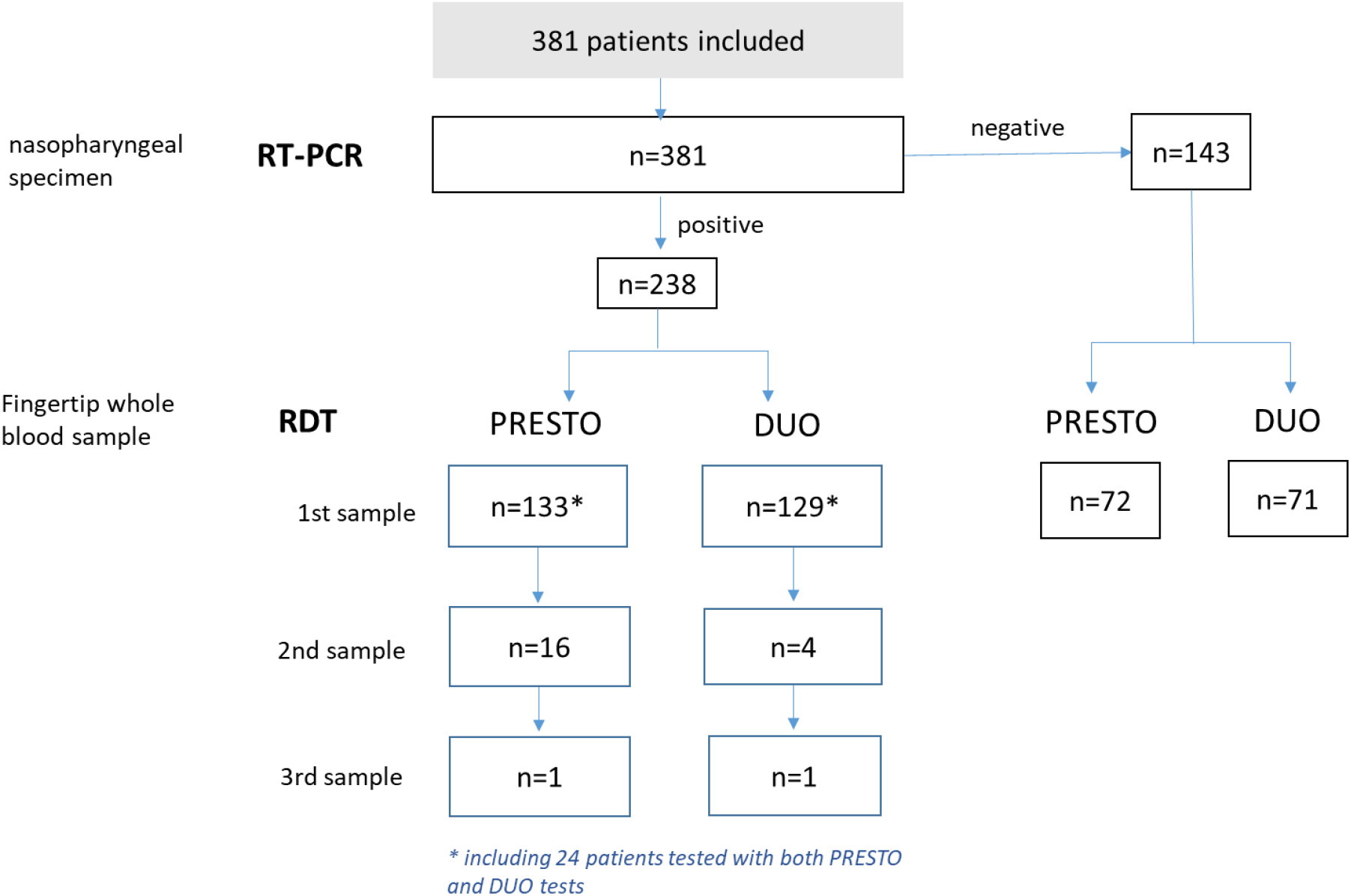
Number of samples screened with RT-PCR and Rapid Diagnostic Tests (RDT)

In the negative RT-PCR subgroup, the mean age was 48.20 years (SD: 17.00; range 19-72), median at 46 years. Among these patients, 72 and 71, respectively, were tested with COVID-PRESTO^®^ and COVID-DUO^®^ tests. All results were negative indicating a specificity of 100% for both RDTs.

In the RT-PCR positive subgroup, the mean age of patients was 53.68 years ± 20.18 (median 54; range 19-96).

For COVID-PRESTO^®^ test, fingertip blood samples were collected from 133 patients, only once (n=133) or at two (n=16) or three different times (n=1). Overall 150 samples used to evaluate the sensitivity of this test. The further the onset of symptoms was from the date of collection, the greater the sensitivity (Table 1): 69.23% [CI95%: 53.58-81.43%] for patients with symptoms that occurred from 11 to 15 days before the date of test and 100% [CI95%: 92.59-100%] in patients who experienced first symptoms more than 15 days before the test. Interestingly, among patient with samples collected at two different times, an elderly woman, 75 years of age, with multiple cancer treated by chemotherapy was negative at Day 15 and positive at Day 19, both for IgM and IgG.

**Table 1.** Evaluation of the sensitivity of the COVID-PRESTO^®^ test.

| Number of days since the onset of symptoms |  |  |  |  |
| --- | --- | --- | --- | --- |
|  | 0-5 days | 6-10 days | 11-15 days | >15 days |
| Positive | 2 | 25 | 27 | 48 |
| Negative | 18 | 18 | 12 | 0 |
| **Sensitivity** | **10.00%** | **58.14%** | **69.23%** | **100%** |
| [CI 95%] | [2.79-30.10%] | [4.33-71.62%] | [53.58-81.43%] | [92.59-100%] |

For COVID-DUO^®^ test, 129 patients were screened with one (n=129), two (n=4) or three samples (n=1) at different times. The sensitivity was assessed based on 134 conducted tests (Table 2). The sensitivity ranged from 35.71% [CI95%: 16.34-61.24%] for patients having experienced their 1^st^ symptoms from 0 to 5 days ago, to 100% [CI95%: 89.85-100%] in patients where symptoms had occurred more than 15 days before the date of tests.

**Table 2.**
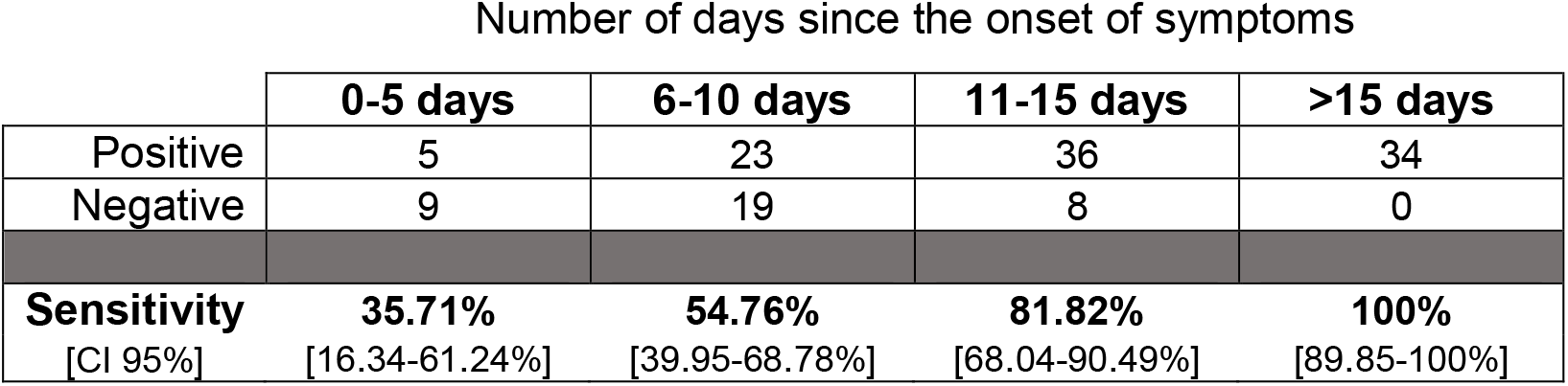
Evaluation of the sensitivity of the COVID-DUO^®^ test.

| Number of days since the onset of symptoms |  |  |  |  |
| --- | --- | --- | --- | --- |
|  | 0-5 days | 6-10 days | 11-15 days | >15 days |
| Positive | 5 | 23 | 36 | 34 |
| Negative | 9 | 19 | 8 | 0 |
| **Sensitivity** | **35.71%** | **54.76%** | **81.82%** | **100%** |
| [CI 95%] | [16.34-61.24%] | [39.95-68.78%] | [68.04-90.49%] | [89.85-100%] |

When considering the distribution of IgM+ and IgG+ patterns among patients with a positive RDT test, the IgM were the first antibodies to be detected and were systematically present in the few positive patients with an onset of symptoms from 0 to 5 days ago (n=2 in COVID-PRESTO^®^ population; n=5 in COVID-DUO^®^). The IgM appeared first and stayed prevalent until 15 days after viral infection then IgG became more frequent (Figs 4 and 5).

**Fig 4.**
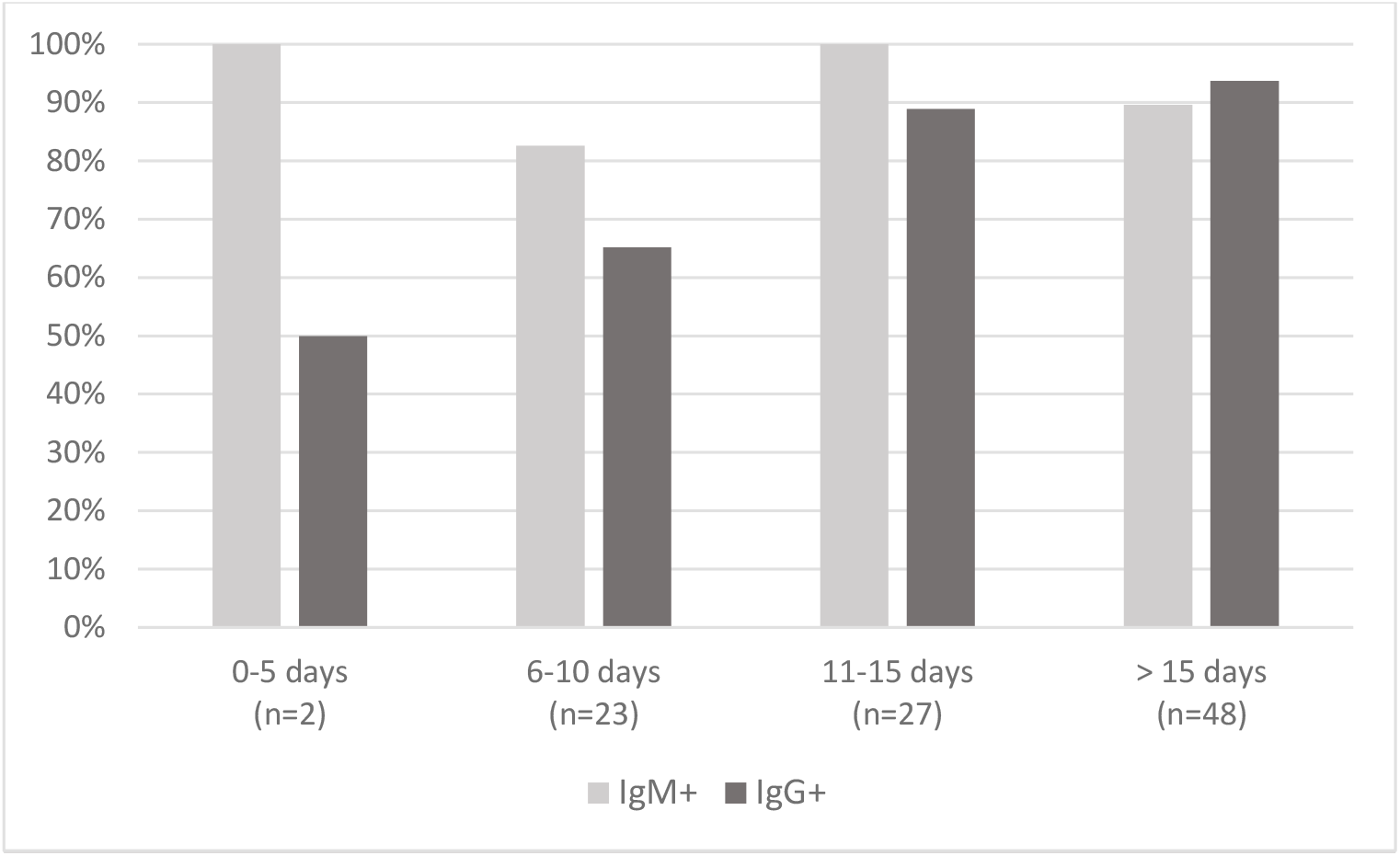
Patients with a positive COVID-PRESTO^®^ test: distribution of IgM+ and IgG+ patterns.

**Fig 5.**
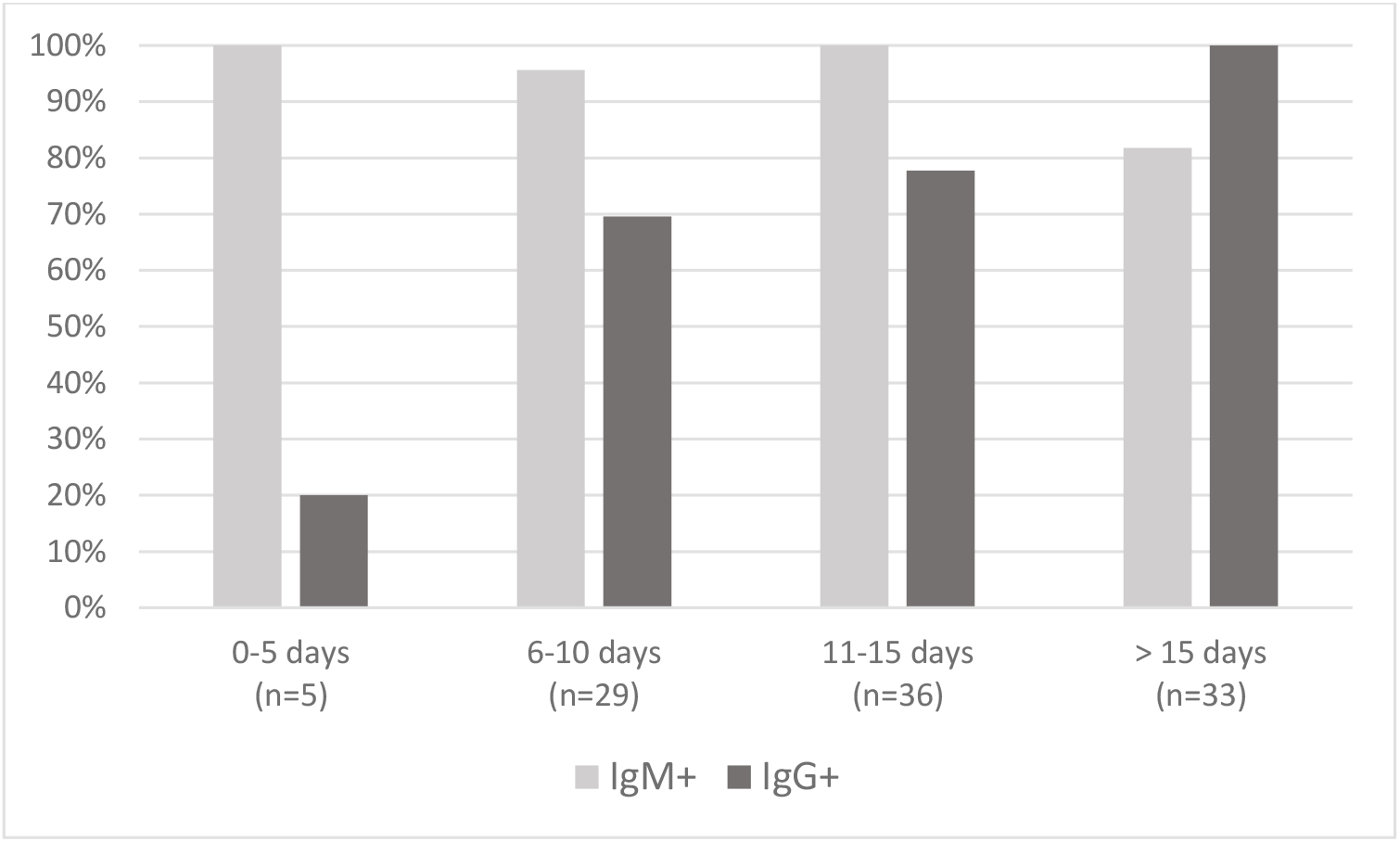
Patients with a positive COVID-DUO^®^ test: distribution of IgM+ and IgG+ patterns.

## DISCUSSION

This real-life study aimed at evaluating the performance of two Rapid Diagnostic Tests (RDT) designed to detect SARS-CoV-2 antibodies IgG and IgM from a fingertip whole blood sample. We investigated the quick detection approach of COVID-PRESTO^®^ and COVID-DUO^®^ in comparison with RT-PCR testing.

The performance analysis was conducted in 381 patients. The results showed that the sensitivity of both RDTs increases with the duration from symptoms onset, reaching 100% in patients experiencing first symptoms of COVID-19 more than 15 days ago. The specificity of both RDTs was found to be 100%, no false positive results having been obtained.

The sensitivity and specificity of such strip assays based on immuno-chromatography have been recently estimated in several studies performed with venous blood samples. In a retrospective study, serum from 179 patients was used to detect SARS-CoV-2 IgG/IgM antibodies [7]. Patients were stratified by the time from symptoms onset to sample collection: 0-7 days, 8-15 days and >15 days. Sensitivities of 18.8%, 100% and 100% were reported, respectively, for the three groups with very few patients (n=8) in the 8-15 days group. The specificity was 77.8%, 50% and 64.3%, respectively, with numerous reported cases of "false positives”. In a second prospective study, the sensitivity of a strip assay investigated in 86 patients was 11.1%, 92.9% and 96.8% at the early stage (1-7 days after onset), intermediate stage (8-14 days after onset), and late stage (more than 15 days), respectively [8]. In another prospective study with 397 PCR confirmed COVID-19 patients and 128 negative patients, the performance of another lateral flow immunoassay test product was evaluated [9]. Overall, the sensitivity was 88.66% and the specificity 90.63%. Although this study was performed with more patients (n=525) than in our study, the evaluation of performance was limited because no information was collected about the period over which each patient had experienced symptoms at the time of blood sample collection. Furthermore, to date, no performance study has been reported based on capillary blood samples.

Although COVID-PRESTO^®^ and COVID-DUO^®^ are only qualitative tests, the reported sensitivities and specificities are closed to those of quantitative assays such as enzyme linked immunosorbent assay (ELISA). Zhao et al. collected blood samples from 173 patients with a confirmed infection with SARS-CoV-2 (acute respiratory infection syndromes and/or abnormalities in chest CT images accompanied by detectable SARS-CoV-2 RNA) at different times after onset of COVID-19: <7 days since onset (early phase), 8-14 days after onset (middle phase) and 13-39 days after onset (later phase) [10]. The detection of IgM and IgG against SARS-CoV-2 in this study was performed using ELISA kits. The sensitivities of IgM assays were 38.3%, 73.3% and 94.3% successively, among samples from patients in early, middle and later phases, respectively. For IgG, the values were 38.3%, 54.1% and 79.8%. Interestingly, the RNA test (RT-PCR on samples from the respiratory tract) had the highest sensitivity (66.7%) in the early phase of illness while RNA was only detectable in 45.5% of samples of day 15-39. From a methodological point of view, the performance study presented here was more robust to that of Zhao et al. because the positive population used as reference to evaluate the sensitivity of RDTs was only based on positive RT-PCR results, and not a mix between syndromes, imagining findings and RNA detection.

The results of the present study highlight two major points. Firstly, as for the assessed RDT, the sensitivity of ELISA tests increases with the duration from symptom onset. Moreover, we showed that direct antibody typing with whole blood is as sensitive as immunoassay performed with serum in a retrospective way. Secondly, these diagnostic tests (either qualitative or quantitative) can help to diagnose a past infection after elimination of the virus by the immune system. Thus, combining RT-PCR and antibody detection allows to largely diagnose COVID-19 people regardless of the delay between infection and diagnosis.

Currently, the extent and the time kinetics of humoral response against SARS-CoV-2 are not known. It is widely accepted that IgM is usually the first responded antibody providing the first line of defense during viral infections, prior to the generation of adaptive, high affinity IgG responses serving as the more robust long term immunity. We were not able to study the humoral response at the individual level because too few patients could have been tested more than once. At the population level, the patterns of IgM/IgG results obtained for positive tests with COVID-DUO^®^ made it possible to perceive the switch between the first production of IgM and the later onset of IgG. This coincided with our observations with the COVID-PRESTO^®^. One of the reasons could lie on the high proportion (90%) of false negative results during the early phase of infection, directly linked to the low titers of antibodies during the first days after infection. Both IgM and IgG titers were found to be low or undetectable 4 days after infection [11] [12]. It was also shown that the presence of antibodies was less than 40% among patients within 1 week since onset, and rapidly increased to 94.3% (IgM) and 79.8% (IgG) from day-15 after onset [10]. The presence of IgM and IgG antibodies against SARS-CoV-2 within 2 weeks from the onset of symptoms was confirmed by others [13] [11]. Recently, in 41 COVID-19 patients confirmed by RT-PCR, it was shown by chemiluminescent immunoassay that the median time of seroconversion was 11 days after disease onset for IgG and 14 days for IgM [14]. The time required to have detectable levels of antibodies explains the poor performance (sensitivity 18.4%) reported for a COVID-19 IgM/IgG Rapid test evaluated in patients admitted to the emergency room [15]. The authors concluded that the Rapid Test lateral Flow Immuno Assay was not recommended for triage of patients with suspected COVID-19. Indeed, COVID-19 cannot be excluded at an early stage when viral serological testing is negative. Although slightly lower than the specificity obtained for COVID-PRESTO^®^ and -DUO^®^, the specificity at early stages was high (91.7%), preventing false positive diagnoses.

This study has several limitations. Firstly, the date of onset of symptoms related to SARS-CoV-2 infection implied recall of facts from memory. This recall bias could lead to some imprecise classification when stratifying the samples by days between onset of symptoms and date of blood samples. Secondly, few patients with a negative serology could have been re-tested with a second blood sample. In these conditions, we were not able to study the dynamics of seroconversion on individual level. Thirdly, there were still negative tests in RT-PCR positive patients up to 15 days after onset. The reasons are multiple and include the relatively low titers of antibody in the early stages of infection as reported by others [16] and the difference in individual immune response antibody production. Lastly, the strength of antibody response depends on several factors, including age, severity of disease, and certain conditions like immunodeficiency disorders. Therefore it would have been interesting to stratify the population depending on immune health. Indeed, we had few subjects with profound immunosuppression who were still negative 15 days after onset. We know, however, that seroconversion could occur later in such patients [17] [18]. Future studies should focus on seroconversion from Day 15 to Day 30 in highly immunocompromised patients infected with COVID-19. However, the highly immunosuppressed patient in this study was well documented to seroconvert between day 15 and day 19, which provides reassurance of the performance of the RDT, even in this population.

Whatever these limitations, COVID-PRESTO^®^ and DUO^®^ RDTs turned out to be very specific (none false positive) and to be sensitive enough after 15 days from onset of symptom. These easy to use IgG/IgM combined test kits are the first ones allowing a screening with capillary blood sample, by typing from a finger prick blood sample. The tests are simple, qualitative, visually interpretable, and give a result within 15 minutes. A positive serology allows to determine whether a person has already been infected by SARS-CoV-2. Serologic tests will be needed to assess the response to vaccine candidates and to map levels of immunity in communities. These rapid tests are particularly interesting for low resource settings such as at the bedside or any other locations where lab tests are less obvious.

### Contributions

TP, JG, GP and LH were responsible for the study design, data interpretation, literature research, and writing of the manuscript. MC, SG, AS, VR, MCC, CK, VR, EL and LC performed the serological testing. TP, MC, CG and LH were responsible for the clinical management, patient recruitment, and data collection. TP, MC JG and LH collected and analyzed the data

## Data Availability

The raw data underlying the results presented in the study are available from Thierry Prazuck, corresponding author.

## Acknowledgments

The authors would like to thank the technical staff of the Department of Infectious diseases for excellent assistance. Furthermore, the authors thank Angèle Guilbot of Clinact, France for providing medical writing support/editorial support in accordance with Good Publication Practice (GPP3) guidelines.

